# Changes in Emergency Department attendances before and after COVID-19 lockdown implementation: a cross sectional study of one urban NHS Hospital Trust

**DOI:** 10.1101/2020.07.20.20157560

**Authors:** K Honeyford, C Coughlan, R Nijman, P Expert, G Burcea, I Maconochie, A Kinderlerer, G S Cooke, C E Costelloe

## Abstract

**Background:** Emergency Department (ED) attendances have fallen across the UK since the ‘lockdown’ introduced on 23^rd^ March 2020 to limit the spread of coronavirus disease 2019 (COVID-19). We hypothesised that reductions would vary by patient age and disease type. We examined pre- and in-lockdown ED attendances for two COVID-19 unrelated diagnoses; one likely to be affected by lockdown measures (gastroenteritis) and one likely to be unaffected (appendicitis).

**Methods:** Retrospective cross-sectional study conducted across two EDs in one London hospital Trust. We compared all adult and paediatric ED attendances, before (January 2020) and during lockdown (March/April 2020). Key patient demographics, method of arrival and discharge location were compared. We used SNOMED codes to define attendances for gastroenteritis and appendicitis.

**Results:** ED attendances fell from 1129 per day before lockdown to 584 in-lockdown; 51.7% of pre-lockdown rates. In-lockdown attendances were lowest for under-18s (16.0% of pre-lockdown). The proportion of patients admitted to hospital increased from 17.3% to 24.0% and the proportion admitted to intensive care increased four-fold. Attendances for gastroenteritis fell from 511 to 103; 20.2% of pre-lockdown rates. Attendances for appendicitis also decreased, from 144 to 41; 28.5% of pre-lockdown rates.

**Conclusion:** ED attendances fell substantially following lockdown implementation. The biggest reduction was for under-18s. We observed reductions in attendances for gastroenteritis and appendicitis. This may reflect lower rates of infectious disease transmission, though the fall in appendicitis-related attendances suggests that behavioural factors are also important. Larger studies are urgently needed to understand changing patterns of ED use and access to emergency care during the COVID-19 pandemic.

**What this paper adds**

- **What is already known on this subject:**
  - **ED attendances have decreased during the COVID-19 associated lockdown.**
  - **Various theories have been suggested for changes in ED attendance, including lower transmission of infectious diseases and patients heeding recommendations from government bodies**.
- **What this study adds:**
  - **Reductions in ED attendances vary by age group and disease type**.
  - **We propose a conceptual casual framework for underlying reasons behind changes in ED attendance in England during the COVID-19 lockdown**.
  - **Following lockdown implementation, we observed a reduction in ED attendances with both infectious and non-infectious diseases. This suggests that reduced transmission of infectious disease is not the only cause of lower overall ED attendances**.

## Introduction

The emergence of COVID-19 and subsequent ‘lockdown’ introduced by the British Government on 23^rd^ March 2020^1^ has had a substantial impact on Emergency Department (ED) attendances. Total ED attendances in England in March 2020 fell by 29.4% year-on-year.^2^ Reasons for this change in ED activity are likely to be multifactorial. To reduce pressure on EDs, patients have been instructed to seek advice from online resources and NHS telephone services. The closure of schools and workplaces is likely to have led to a reduction in the spread of infectious diseases.^2^ Reductions in organised sports and recreational activity will be associated with reductions in physical injuries.^3^ It is suggested that reductions in children attending EDs reflect parents’ concerns about acquiring infection in hospital settings.^4^

We hypothesised that the impact of the nationwide lockdown on ED attendance would vary by patient demographics and clinical reason for attendance. We have proposed causal pathways leading to changes in ED attendances and hospital admissions. These are summarised in Figure 1. Using a snapshot of ED data, we examined the number of pre- and in-lockdown ED attendances for two COVID-19 unrelated diagnoses:

**Figure 1.**
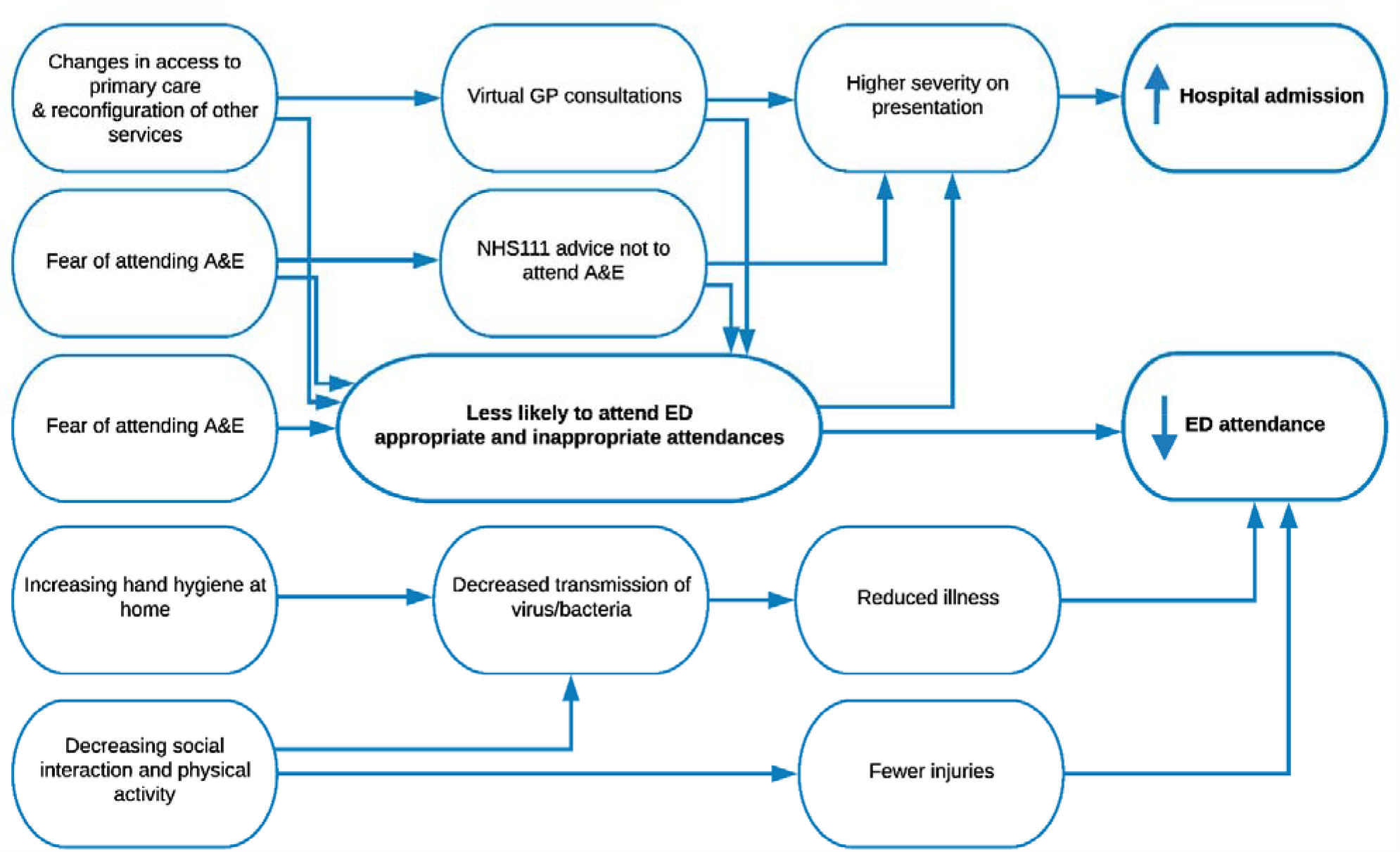
Conceptual framework for changes in ED attendance associated with ‘lockdown’ for conditions unrelated to COVID-19.

- Gastroenteritis – an infectious disease which we would expect to be affected by lockdown measures.
- Appendicitis – an acute disorder which we would expect to be largely unaffected by lockdown measures.

## Methods

ED attendance information was provided by the NHS Trust data warehouse team. Information included patient information: age, gender, ethnicity, residential partial postcode, arrival mode at ED, destination at discharge and SNOMED^5^ diagnostic codes.

We selected a four-week period (6^th^ January 2020 to 2^nd^ February 2020) as the pre COVID-19 phase and a four-week period (23^nd^ March 2020 to 19^th^ April 2020) as the in-lockdown period. We included all ED attendances (children and adults).

We compared ED attendances based on patient demographics and we compared attendances for gastroenteritis and appendicitis, in total and by age, to assess examples of one diagnosis likely to be affected and one diagnosis unlikely to be affected by lockdown measures. Differences were assessed using Chi-Squared test. Analysis was done in R version 3.60. This study was granted service evaluation approval through Imperial College London NHS Trust (Ref:228). Patients or the public were not involved in the design, or conduct, or reporting, or dissemination plans of our research.

## Results

There were 31,624 ED attendances in the pre-lockdown period and 16,355 in-lockdown; a reduction from 1129 attendances a day pre-lockdown, to 584 a day in-lockdown. Arrivals in ambulances accounted for 61.2% attendances pre- and 51.7% in-lockdown. As a proportion of pre-lockdown attendances, in-lockdown attendances were lowest for under-18s (16.0%) and highest for patients aged 40-60 (76.7%). Male and Asian patients made up a higher proportion of in-lockdown than pre-lockdown attendances. This was also true for patients from postcodes considered the primary catchment for the Trust (77% pre-lockdown and 80% in lockdown). Pre-lockdown, 17.5% of ED attendances resulted in admission to inpatient wards or intensive care units (ICUs), compared to 24.4% in-lockdown. Following lockdown implementation, 4% of admitted patients were admitted directly to ICU, compared to 1% pre-lockdown. Results are summarised in Table 1.

**Table 1.**
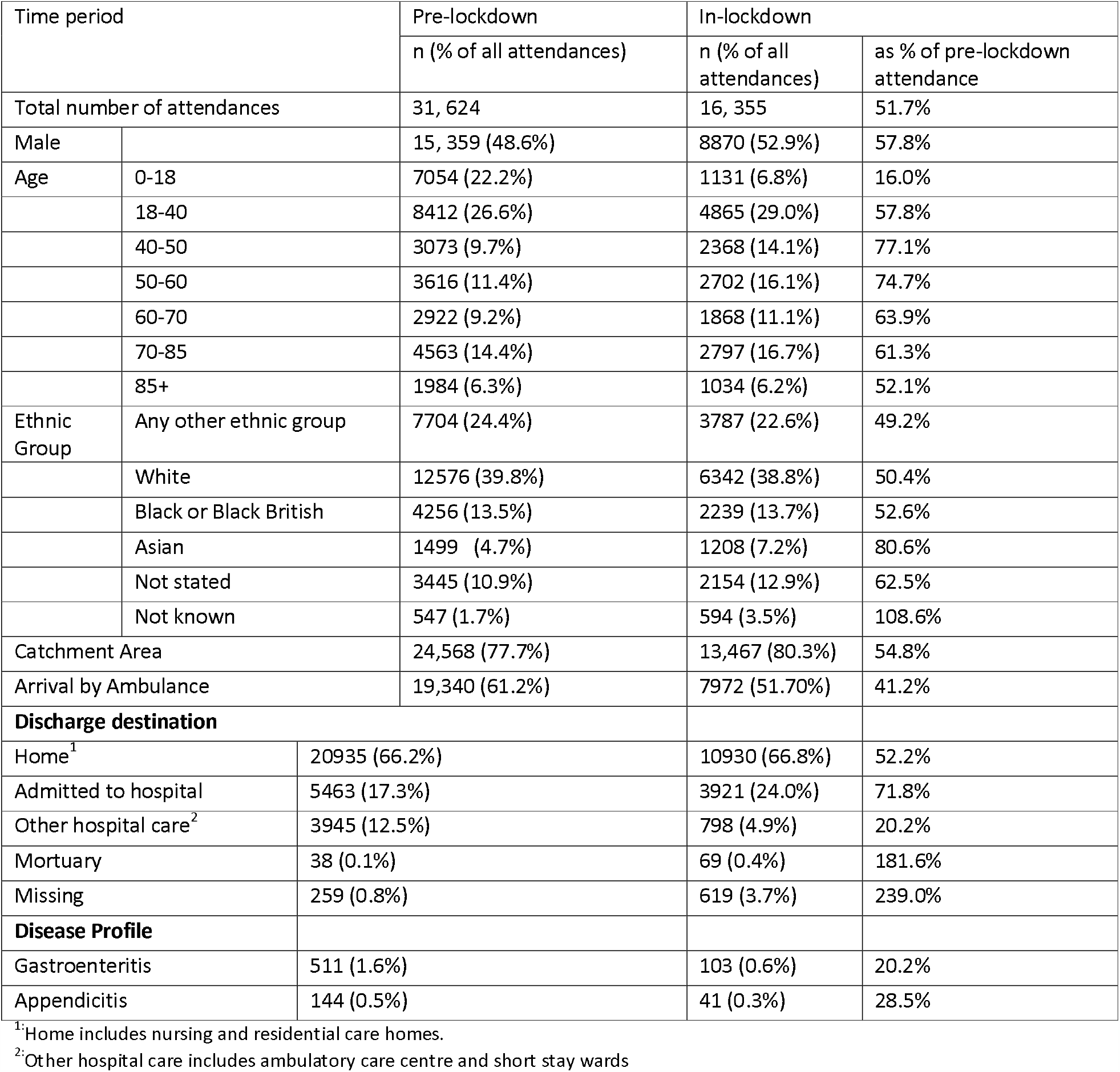
Characteristics of patients attending two EDs in one hospital trust in North West London, p<0.0001 for all pre vs in-lockdown comparisons.

### Gastroenteritis and appendicitis attendances pre- and in-lockdown

Pre-lockdown, there were 511 attendances with a gastroenteritis code, 1.62% of all attendances, compared to 103 attendances in-lockdown, 0.61% of the total. Total ED attendances with an appendicitis code also decreased over the study period, from 144 (0.46% total) to 41 (0.24% total). Attendances for gastroenteritis in-lockdown were 20.2% of pre-lockdown, compared to 28.5% for appendicitis. While a similar proportion of patients with gastroenteritis were directly discharged home in both time periods, we observed a three-fold increase in discharge rates among patients with appendicitis following lockdown implementation.

Changes in attendances for both diseases varied with age (Figure 2). We observed the most significant reduction in attendances with gastroenteritis amongst children and young people and patients aged over 60.

**Figure 2.**
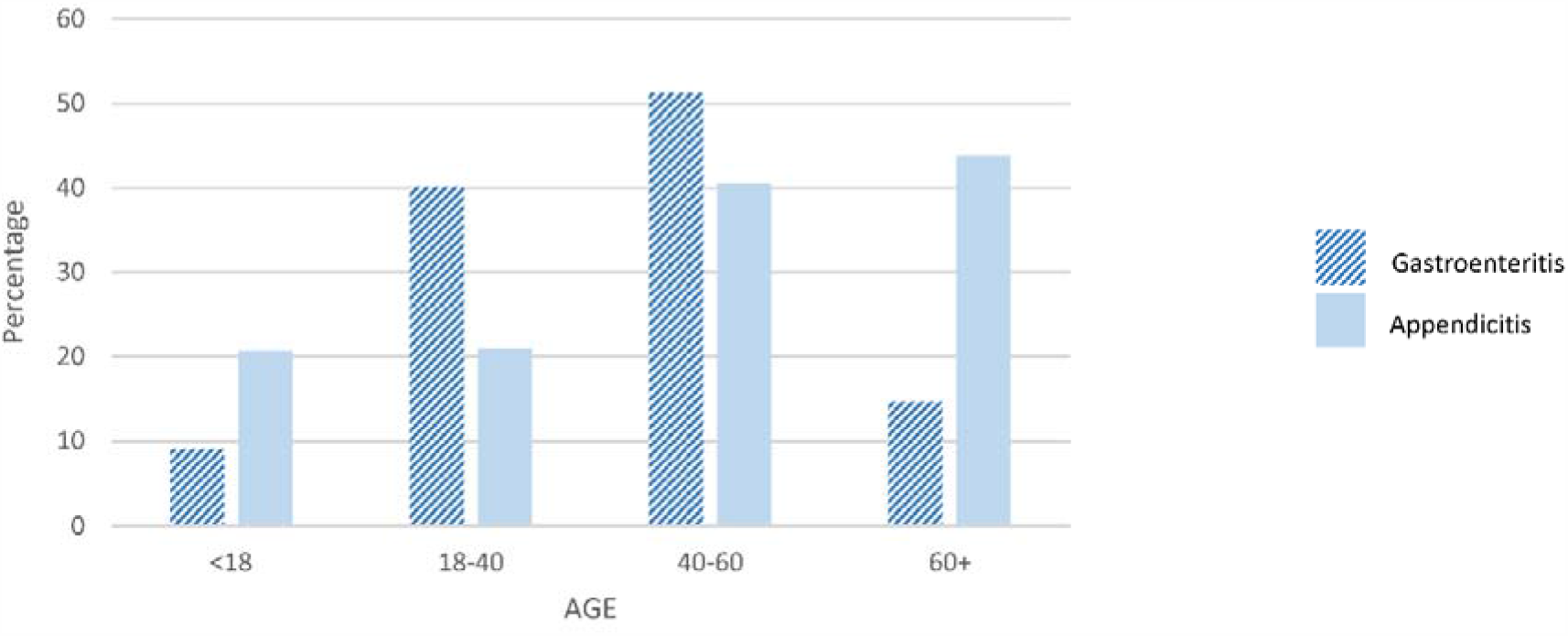
ED attendances for gastroenteritis and appendicitis, by age, post-lockdown-implementation as a percentage of pre-lockdown attendances.

## Discussion

In line with national data^2^, we have found that overall ED attendances have almost halved since the introduction of lockdown. Similar to other reports^6^ the impact of the lockdown on ED attendance rate was greatest in the under-18’s, suggesting changes in parental health-seeking behaviour.

Following lockdown implementation, a higher proportion of ED patients required hospital admission and there was a four-fold increase in the proportion admitted directly to ICU. These changes may reflect patients attending with more serious conditions, severe COVID-19 and/or the increase in ICU capacity.

We hypothesised a reduction in gastroenteritis-related attendances following lockdown implementation due to reduced interpersonal contact and spread of infectious diseases. The results show that attendances in-lockdown fell to one fifth of pre-lockdown rates. A reduction in ED attendances for specific infectious diseases has been described in England^2^ and Italy.^7^ However, the decrease in attendances for appendicitis suggests that reduced transmission alone cannot explain the reduction in ED attendances seen after lockdown.

Not all of the differences we have reported are attributable to lockdown Seasonal variations are seen in a range of infectious diseases, including gastroenteritis. Nationally published data for this hospital Trust suggests April attendances in 2019 were only 5% lower than January 2019.^8^ Patients with COVID-19 may present with diarrhoea and vomiting^9^ and this may confound gastroenteritis coding. Some attendances will be due to COVID-19; we did not exclude these as it’s impossible to know patients’ motivation for attending the ED and testing policy evolved over time. Treatment pathways have changed during the pandemic as hospitals have sought to minimize non-emergency surgery. This includes an increase in the management of uncomplicated appendicitis with oral antibiotics possibly explaining the fall in appendicitis-related admissions.^10^

We have developed a conceptual causal framework proposing various factors which may lead to lower ED attendances during the COVID-19 pandemic. These factors include deterred care seeking due to fears of acquiring infection in hospital settings and patients seeking health advice from other services. Future studies, using larger, more generalisable data must aim to untangle the relative contributions of these different factors and ensure that sick patients have timely and equitable access to Emergency Care.

## Data Availability

Data is available through: The Imperial College Healthcare NHS Trust (ICHT) Clinical Analysis, Research and Evaluation (iCARE) platform.
Requests to access ICHNT and NWL's WSIC COVID-19 de-identified datasets on iCARE are reviewed and approved by the COVID-19 NWL Data Prioritisation Group. This group meets weekly to review both operational and research related requests to access the de-identified data for COVID-19 related projects and is chaired by the Sector wide Caldicott guardian. The COVID-19 Data Prioritisation Group also maintains oversight of dataset development and ensures clinical oversight of all research. To be allowed access, a researcher must be sponsored by a senior clinician practicing at one of the relevant data controllers.

https://imperialcollegehealthpartners.com/covid-19-data-prioritisation-group/

## AUTHOR CONTRIBUTIONS

KH, CC, RN, ChC and PE conceived the study and developed the protocol. KH conducted the statistical analysis, which was reviewed and improved by PE, GB and CC. GSC, RN, ChC, IM and AK provided ongoing feedback on results and their interpretation. KH wrote the first draft of the manuscript. All authors reviewed and contributed to the final draft of the manuscript. CC is the guarantor. KH attests that all listed authors meet authorship criteria and that no others meeting the criteria have been omitted. The views expressed are those of the author(s) and not necessarily those of the National Health Service, National Institute for Health Research, or the Department of Health and Social Care.

## ACKNOWLEDGMENTS

This research was funded by the NIHR Imperial Biomedical Research Centre (BRC). The research was conducted using National Institute of Health Research Health Informatics Collaborative data resources. The views expressed are those of the authors and not necessarily those of the NIHR or the Department of Health and Social Care.

Imperial College London is grateful for support from the NW London NIHR Applied Research Collaboration. The views expressed in this publication are those of the authors and not necessarily those of the NIHR or the Department of Health and Social Care

## References

1 https://www.parliament.uk/business/news/2020/march/update-on-coronavirus-24-march-2020

2 Kelly E & Firth Z How is COVID-19 changing the use of emergency care? The Health Foundation. 15 May 2020. Available from: www.health.org.uk/news-and-comment/charts-and-infographics/how-is-covid-19-changing-the-use-of-emergency-care. Accessed: 5th June 2020.

3 Buddhdev P, Gille H and Ibrahin Y The paediatric burden of UK lockdown – early results in the COVID-19 era. British Orthopaedic Association (2020). Available at: https://www.boa.ac.uk/policy-engagement/journal-of-trauma-orthopaedics/journal-of-trauma-orthopaedics-and-coronavirus/the-paediatric-trauma-burden-of-uk-lockdown-early.html Accessed: 9th June 2020

4 Wong LE, Hawkins JE, Langness S et al. Where Are All the Patients? Addressing Covid-19 Fear to Encourage Sick Patients to Seek Emergency Care | Catalyst non-issue content [Internet]. Catalyst.nejm.org. 2020. Available from: https://catalyst.nejm.org/doi/full/10.1056/CAT.20.0193 Accessed 9th June 2020

5 NHS Digital SNOMED CT 2020 Available from:https://digital.nhs.uk/services/terminology-and-classifications/snomed-ct Accessed 9th June 2020

6 Isba R, Edge R, Jenner R, et al Where have all the children gone? Decreases in paediatric emergency department attendances at the start of the COVID-19 pandemic of 2020 Archives of Disease in Childhood Published Online First: 06 May 2020. doi: 10.1136/archdischild-2020-319385

7 Angoulvant F, Ouldali N, Dawei Yang D et al COVID-19 Pandemic: Impact Caused by School Closure and National Lockdown on Pediatric Visits and Admissions for Viral and Non-Viral Infections, a Time Series Analysis. 202 Clinical Infectious Diseases. https://doi.org/10.1093/cid/ciaa710.

8 NHS England. A&E Attendances 2020 Available from: www.england.nhs.uk/statistics/statistical-work-areas/ae-waiting-times-and-activity/ Accessed 5th June 2020

9 Jin X, Lian J, Hu J, et al. Epidemiological, clinical and virological characteristics of 74 cases of coronavirus-infected disease 2019 (COVID-19) with gastrointestinal symptoms. Gut 2020 https://gut.bmj.com/content/69/6/1002

10 Collard M, Lakkis Z, Loriau J, et al J Visc Surg. Antibiotics alone as an alternative to appendectomy for uncomplicated acute appendicitis in adults: Changes in treatment modalities related to the COVID-19 health crisis 2020 Apr 24 doi: 10.1016/j.jviscsurg.2020.04.014 [Epub ahead of print]

